# Characterization of the somatic landscape and transcriptional profile of breast tumors from 748 Hispanic/Latina women in California

**DOI:** 10.64898/2026.02.13.26346286

**Authors:** Yuan Chun Ding, Rosalyn W. Sayaman, Denise Wolf, Joanne Mortimer, Shu Tao, Allen Mao, Laura Fejerman, Stephen B. Gruber, Stephen Francis, Susan L. Neuhausen, Elad Ziv

## Abstract

Somatic mutations and the tumor immune microenvironment (TIME) in breast tumors are important predictors of treatment response and survival, yet data for Hispanic/Latina (H/L) women are limited. Here we analyzed whole exome sequencing data from tumor/normal pairs and RNAseq data from 748 H/L women and 388 non-Hispanic White (NHW) women. Despite very similar somatic mutational and copy number profiles, we found significant differences in TIME ecotypes. TIME ecotypes CE9 and CE10, characterized by increased lymphocyte infiltration and more favorable prognosis, were more common among women with higher Indigenous American (IA) ancestry. A germline *APOBEC3A/B* copy-number deletion, significantly more common in H/L than NHW women, was associated with APOBEC mutational signatures and with CE10 ecotype. The *APOBEC3A/B* copy-number deletion was associated with overexpression of transposable elements (TEs) and TEs were strong predictors of CE10 ecotype. This suggests that the *APOBEC3A/B* and CE10 association may be driven via the disinhibition of these TEs. Overall, these results demonstrate the utility of characterizing previously understudied populations to infer new mechanisms of cancer immune responses.

## INTRODUCTION

Somatic mutations and copy number aberrations are important predictors of outcomes among women with breast cancer ^1,2^. Increasingly, somatic profiles are being used to identify effective therapies. Tumors with amplifications of *ERBB2* can be targeted by both antibody and small molecules directed at this receptor ^3^. Tumors with activating mutations in *PIK3CA* can be treated with PI3Kinhibitors with response dependent on the mutation ^3–5^. Tumors in women with *BRCA1, BRCA2* and *PALB2* pathogenic variants respond to PARP inhibitors ^6–8^ and platinum-based chemotherapy ^9^. In addition, the immune tumor microenvironment (TME) is an important predictor of treatment outcomes, and, in particular, of treatment with immune checkpoint inhibitors ^10–13^.

The majority of publicly available datasets of breast tumors are from women of European ancestry ^14^. Analyses of The Cancer Genome Atlas (TCGA) suggest that there may be differential effects of some somatic events and mutational signatures by ancestry ^15,16^. For example, a higher proportion of breast tumors in women of African ancestry are triple negative for hormone receptor and human epidermal growth factor receptor 2 (HER2) status (triple negative), have fewer mutations in *PIK3CA,* are homologous recombination deficient (HRD), and have more TP53 mutations compared with breast tumors from women of European ancestry ^17–19^. East Asian women and Hispanic/Latino (H/L) women with high Indigenous American (IA) ancestry are more likely to develop tumors with HER2 amplification ^20–22^ and East Asian women are more likely to develop tumors with *EGFR* mutations ^23^.

H/L individuals constitute ∼18% of the US population and are of diverse genetic ancestry with varying proportions of European, IA, Asian, and African influence ^24–29^. We ^30^ and others ^31^ previously performed tumor/germline whole exome sequencing and RNAseq profiling of tumors from H/L women, but these studies had relatively modest sample sizes and were not well-powered to detect rare events in this population. Here we comprehensively analyze tumor-normal whole exome sequencing and RNAseq data from 748 H/L women and 388 non-Hispanic White (NHW) women and evaluate both their somatic mutational landscape and immune TME.

## RESULTS

### Clinical, demographic, and ancestry features

We combined germline genetic, tumor genomic, and tumor transcriptomics to comprehensively characterize breast tumors from H/L and NHW women (Figure 1). We compared data from 436 H/L and 388 NHW women in the Implementing Next-generation Sequencing for Precision Intervention and Risk Evaluation (INSPIRE) study. We also included data from 312 H/L women in the California Latina Breast Cancer Study (CLBCS) for a combined data set of 748 H/L women to increase power for analyses focused on H/L women and IA ancestry. We inferred genetic ancestry using ADMIXTURE analysis and included reference ancestry groups from the Human Genome Diversity Project (HGDP). The majority (75%) of NHW from the INSPIRE study had high (> 80%) European ancestry (Figure 2A), whereas H/L from both the INSPIRE (Figure 2B) and CLBCS studies (Figure 2C) were of predominantly mixed European and IA ancestry with an average of 39% IA ancestry varying widely from <1% to > 99% at the extremes. Using principal component analysis, we examined genetic similarity among individuals from the INSPIRE study. As expected, H/L mostly clustered between the European and IA HGDP reference populations (Figure 2D).

**Figure 1.**
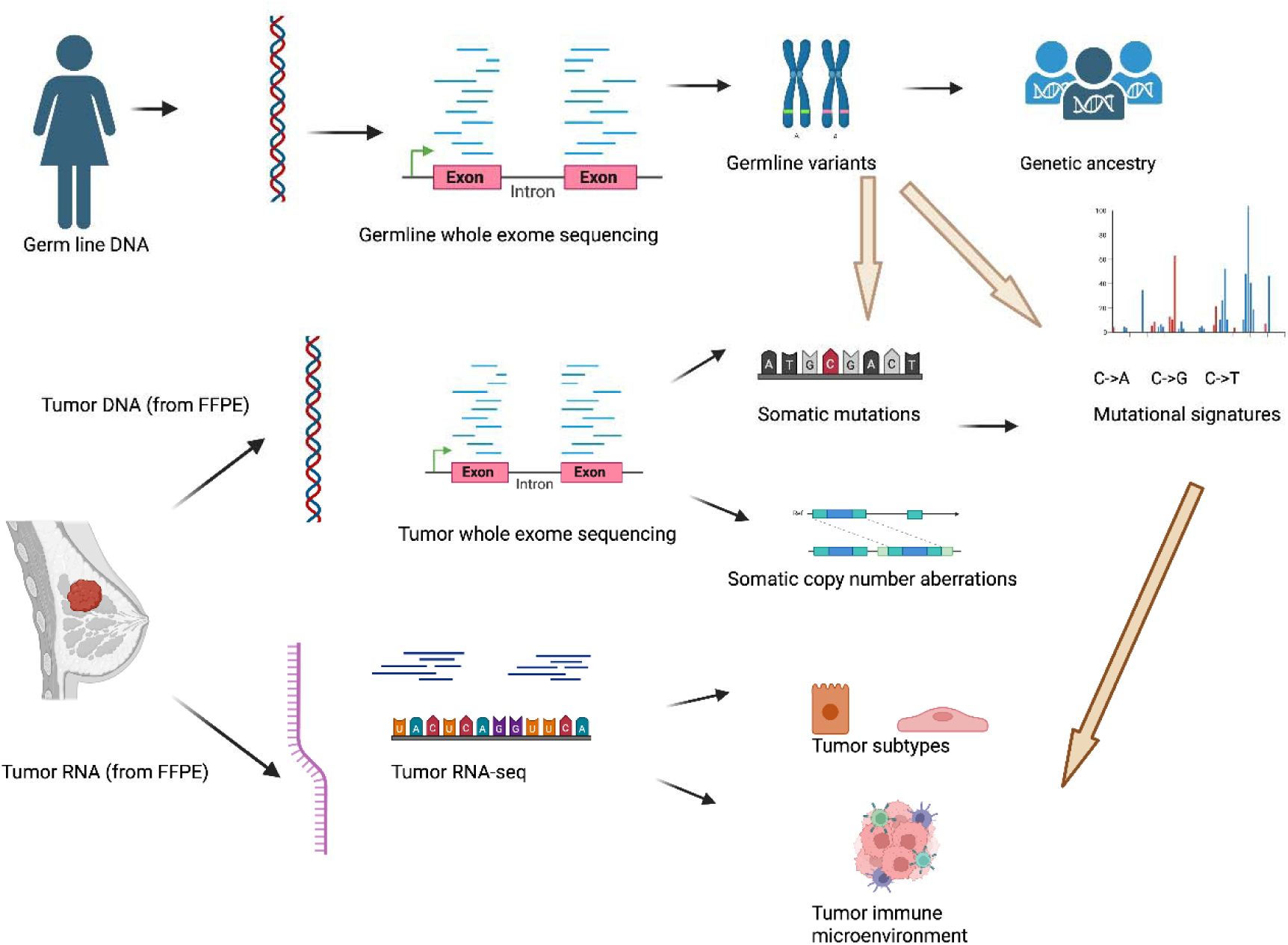
Multi-omic characterization of breast tumors in Hispanic/Latina (H/L) and Non-Hispanic White women. Germline DNA was subjected to whole-exome sequencing to infer germline genetic variation and genetic ancestry. Tumor DNA extracted from formalin-fixed paraffin-embedded (FFPE) tissue underwent whole-exome sequencing to identify somatic single-nucleotide variants and small insertions/deletions, as well as somatic copy number aberrations. Tumor RNA isolated from FFPE samples was analyzed by RNA sequencing to characterize gene expression profiles, tumor intrinsic subtypes, and features of the tumor immune microenvironment. Somatic mutation data were further used to derive mutational signatures. Integrated analyses across germline, somatic, and transcriptomic data enabled comprehensive characterization of breast tumor biology across ancestry groups.

**Figure 2.**
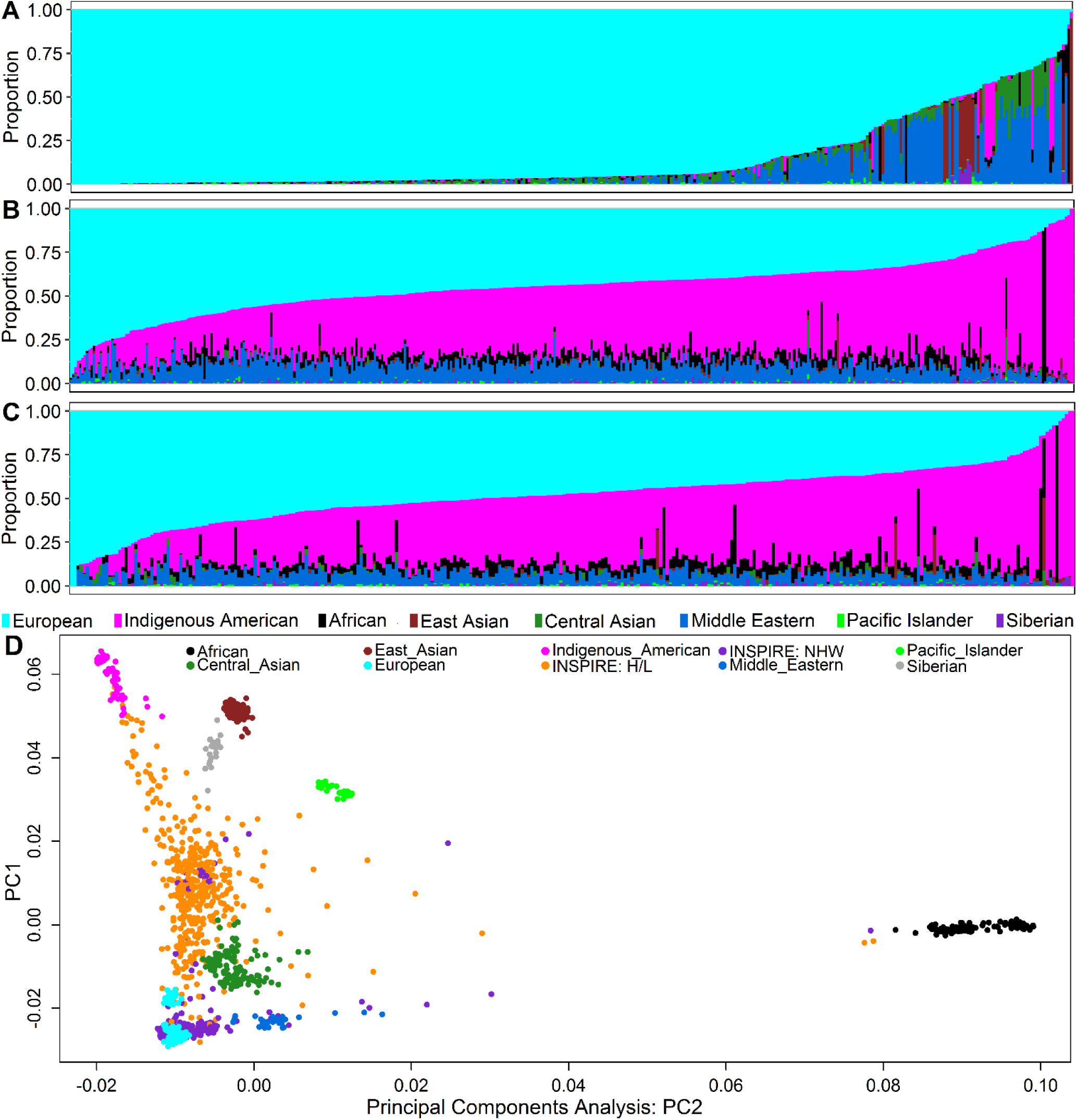
Ancestry inference for patients. **(A,B,C)**: stacked bar graph of genetic ancestry proportions of 8 ethnic reference groups: (**A)** 388 Non-Hispanic whites (NHW) from the INSPIRE study; (**B**) 436 Hispanic/Latinas (H/L) from the INSPIRE study; (**C)** 312 H/L from the CLBCS; each vertical bar represents stacked fractions or proportions of ancestry from one individual estimated using the Admixture program. Ancestry was assigned for each individual as a fraction of European (cyan), Indigenous American (magenta), African (black), East Asian (brown), Central Asian (forest green), Middle Eastern (blue), Pacific Islander (light green), and Siberian (purple). (**D)** Scatterplots of Principal Component 1 (PC1, y-axis) versus PC2 (x-axis) from principal components analysis to show 8 reference ancestry groups and 824 samples from INSPIRE study. Each dot represents the results from one individual: 8 ethnic reference groups from the Human Genome Diversity project (HGDP) including European (cyan), Indigenous American (magenta), African (black), East Asian (brown), Central Asian (forest green), Middle Eastern (blue), Pacific Islander (light green), Siberian (gray) and 824 test samples from the INSPIRE study [H/L (orange) and NHW (purple).

Among breast cancer patients enrolled in the INSPIRE study, H/L women (n = 436) had a mean diagnosis age of 50.5 years (median of 50 years) and were significantly younger than the NHW women (n = 388) who had a mean diagnosis age of 54.1 years (median of 55 years) (P < 0.000001) (Table 1). Women in the CLBCS study (n = 312) were younger than those in the INSPIRE study (mean of 48.5 years and median of 48 years) (Table 1). In the INSPIRE study, H/L women had significantly fewer stage I and more stage II/III breast cancers compared to NHW women (P < 0.00001). After adjusting for age at diagnosis, H/L women remained significantly more likely to present with stage II and III versus stage I breast cancer (both P < 0.001). The distribution of tumor stage (I, II, and III) in the H/L tumors was similar in the INSPIRE and CLBCS datasets. More tumors among H/L woman were estrogen receptor (ER)-negative (22.2%) compared to NHW (16.9%) in the INSPIRE study (p = 0.07) (Table 1). There were no significant differences in progesterone receptor (PR) and HER2 status. We also observed a trend towards a higher proportion of triple negative tumors with 14.2% in H/L in comparison to 10.8% in NHW (p = 0.1, Table S1). For PAM50 subtypes, H/L had significantly higher rates of Basal-like and HER2-enriched subtypes than NHW in the INSPIRE study and a lower rate of Luminal A subtype (p = 0.05). Because the CLBCS study excluded breast cancer patients who had received neoadjuvant treatment, there were lower rates of ER-negative, PR-negative, and triple negative breast cancers than was observed in the unselected H/L cases in the INSPIRE study; this also was reflected in lower rates of Basal and HER2-enriched subtypes and higher Luminal A subtype among the three groups.

**Table 1.**
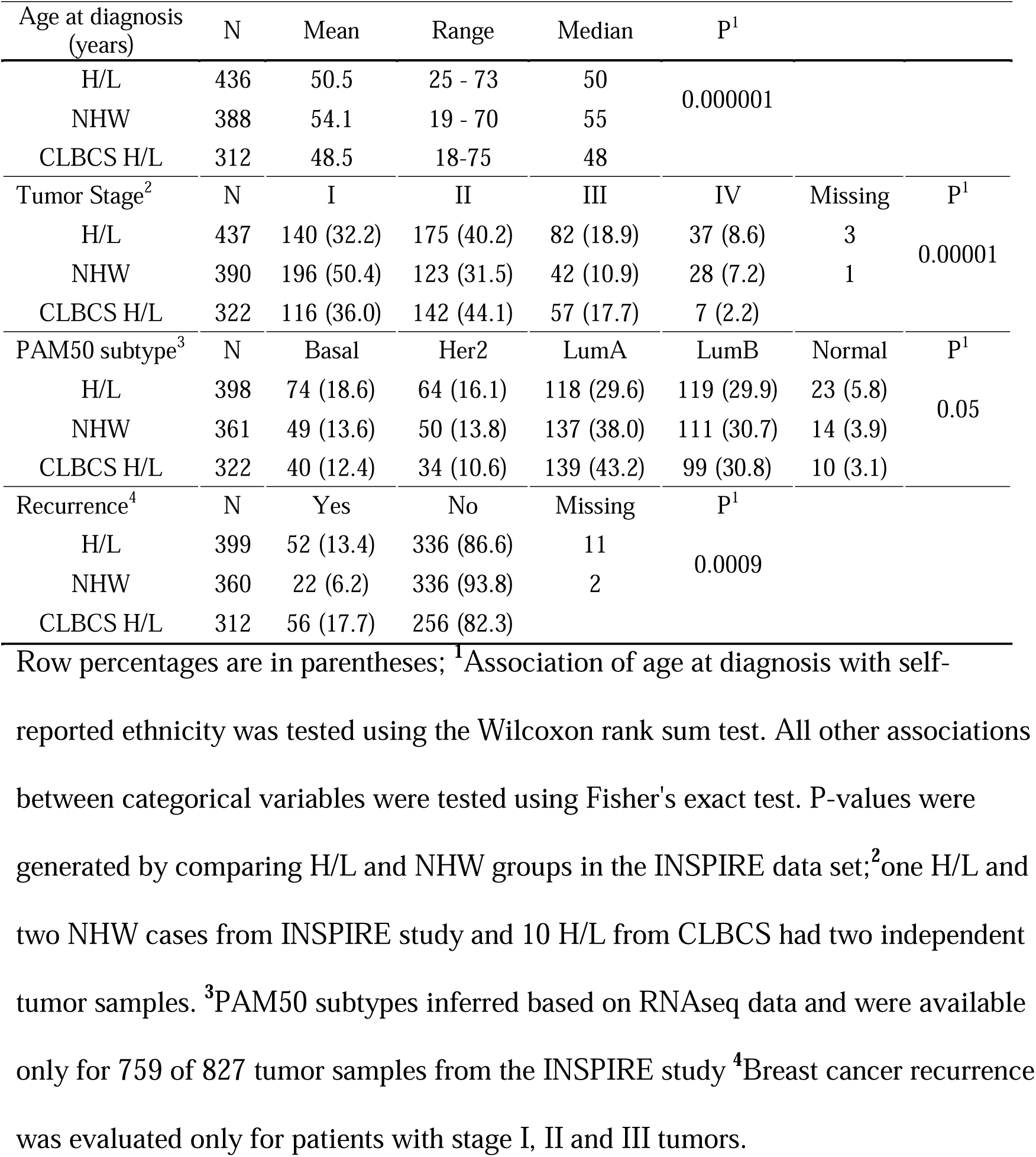
Patient and tumor characteristics of 436 Hispanic/Latinas (H/L) and 388 Non-Hispanic Whites (NHW) from the INSPIRE study and 312 H/L from CLBCS.

In the INSPIRE study, H/L had two-fold higher breast cancer recurrence than NHW (13.4% versus 6.2%); this association (P < 0.001) remained significant in the multivariable logistic regression analysis adjusting for age at breast cancer diagnosis, tumor stage and PAM50 subtype (p = 0.026). Among women in the CLBCS, 56 of 312 (17.7%) experienced recurrence, a higher rate than in the INSPIRE study, likely attributable to the longer mean follow-up in CLBCS (6.3 vs. 2.6 years).

### Somatic mutations

We identified a total of 22 significantly mutated genes (FDR < 0.05) (Figure 3A) in the INSPIRE study. Fourteen genes were significantly mutated in both H/L and NHW, three were significant only in H/L (*CTCF, HS6ST1*, *FOXA1)* and five genes were significant only in NHW (*CDKN1B, PRRT2, MUC4, NCOR1, ARID1A*) at FDR < 0.05. Of those 22 genes, 12 were significantly mutated in CLBCS (MutSigCV q value < 0.05) and 20 of 22 genes had at least one non-silent mutation (Table S2). We combined data from the H/L cohorts to test whether somatic mutations in the 20 genes common to both studies were associated with IA ancestry. None of the genes were statistically significantly associated with ancestry after adjustment for multiple hypothesis testing (Table S3).

**Figure 3.**
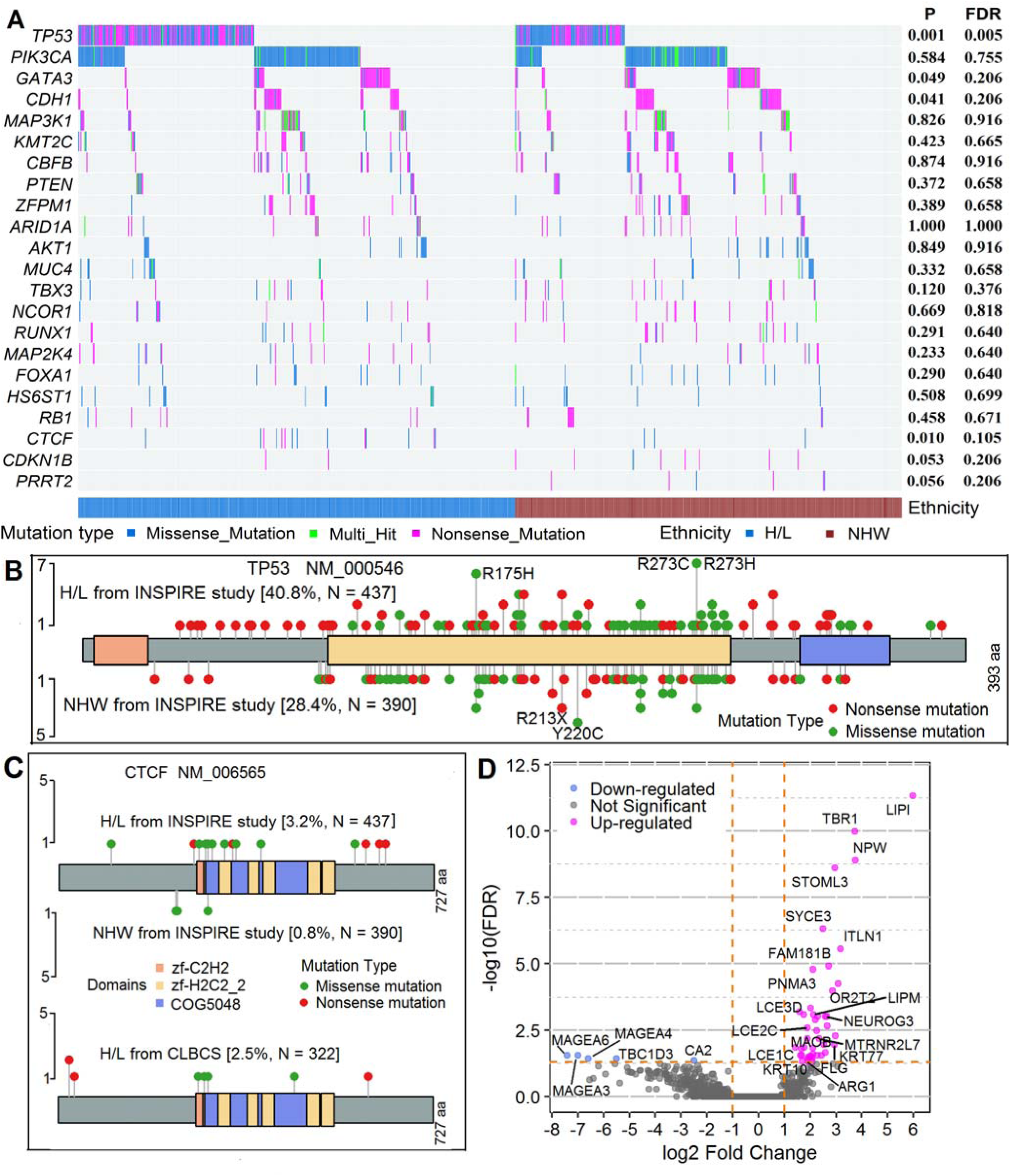
Oncoplot and lollipop plots to display somatic mutations. (**A)** Oncoplot of somatic mutations in 22 cancer driver genes in 437 Hispanic/Latina (H/L) and 390 Non-Hispanic White (NHW) from the INSPIRE study. For the mutation type, in-frame insertion and in-frame deletion mutations are grouped into the “missense mutation”; frame-shift, nonsense, non-stop, splice-site, and translation-start-site mutations are grouped as “truncating mutation”. For each gene, Fisher’s exact test was used to test frequency difference in somatic mutations between H/L and NHW; p values and false discovery rates (FDR) are shown on the right side of the oncoplot. (**B)** Lollipop plot of *TP53* somatic mutations from H/L and NHW breast tumors in the INSPIRE study. (**C**) Lollipop plots of somativ *CTCF* mutations from H/L and NHW tumors in the INSPIRE study and H/L tumors in CLBCS. (**D**)Volcano plot of differentially expressed genes asociated with somatic mutations in *CTCF* in 582 ER positive samples.

Non-silent mutations in *TP53* were significantly more common in H/L (40.8% frequency of mutation) than in NHW in INSPIRE (28.4%) (p = 0.00024, FDR = 0.005, Figure 3A and 3B). However, the frequency of non-silent *TP53* mutations in CLBCS was only 27.4%, similar to NHW (28.4%) in the INSPIRE study. *TP53* mutations were significantly associated with non-Luminal A PAM50 subtypes (Tables S4 and S6), with mutations highly enriched in the basal subtype (p < 1 ×10^−16^ in INSPIRE and p = 1.34 ×10^−14^ in CLBCS), followed by HER2 subtype (p = 2.36 ×10^−16^ in INSPIRE and p = 3.68 ×10^−11^ in CLBCS) and Luminal B subtype (p = 5.56 ×10^−7^ in INSPIRE and p = 1.34 ×10^−6^ in CLBCS), similar to what has been observed previously ^32^. In multivariable models adjusted for PAM50 subtype and stage, the association with H/L ethnicity was no longer significant and there was a strong association with PAM50 subtype. (Table S6).

Mutations in *CTCF* were more frequent in H/L (3.2%) than in NHW (0.8%) in the INSPIRE study but did not meet the FDR < 0.05 criterion (nominal p = 0.0095; FDR = 0.10; Figure 3C). Similar to the frequency in H/L tumors in the INSPIRE study, 2.5% of CLBCS tumors had non-silent mutations in *CTCF*. *CTCF* mutations were enriched only in ER-positive tumors, Luminal B and Luminal A subtypes (absent in Basal and HER2 subtypes) and were mutually exclusive with tumor suppressor gene *TP53* mutations (Table S4). ^33,34^. CTCF affects genome organization and regulates chromatin accessibility and transcription for many genes ^33,34^. Therefore, we examined the association between *CTCF* somatic mutational status and gene expression in our combined dataset of 542 ER+ tumor samples from H/L women. We tested whether the differential expression fold changes were significantly greater than 1.5 and found 5 genes had lower expression and 44 genes (FDR < 0.05) had higher expression in women with *CTCF* mutations after adjusting for tumor purity, datasets, tumor stage, age at breast cancer diagnosis, PAM50 subtype and IA ancestry (Figure 3D). Gene set enrichment analysis identified upregulated epithelial differentiation genes and downregulated histone deacetylase pathway genes (Table S5).

### Somatic mutation signatures

Using the Sigprofile assignment tool, Catalogue of Somatic Mutations in Cancer (COSMIC) single base substitution (SBS) mutation signatures were assigned to 762 tumor samples with at least 10 single-base-pair mutations in the INSPIRE study. COSMIC mutational signatures provide a profile of the etiology and evolution of the biology of tumors. Ten of 26 signatures (SBS1, 2, 3, 5, 6, 7, 10, 13, 15, 16) were detected in at least 5% of samples in either the H/L or NHW groups (Table S7). SBS5, a clock-like signature, was more common in tumors from NHW compared to tumors from H/L (p < 0.05 by both Wilcoxon test and multiple regression) (Table S7). We also found that SBS2, one of two signatures of APOBEC activity, showed a trend towards being more common in tumors from H/L than tumors from NHW (p = 0.054 by Wilcoxon test and p = 0.128 by multiple regression), which may be attributed to a higher frequency of the germline *APOBEC3A/B* deletions in H/L than in NHW women ^30^.

### Somatic copy number

In the INSPIRE study, an integrated analysis of copy number using FACETS and GISTIC2 identified statistically significant (q < 0.05) focal gains (n = 93) and focal losses (n = 122). Of the gains, 19 were H/L-specific, 24 NHW-specific, and 50 shared; of the losses, 16 were H/L-specific, 19 NHW-specific, and 87 shared (Figure S1). Integrating RNAseq expression with gene-level GISTIC2-thresholded copy-number score (−2 deep loss, −1 shallow loss, 0 neutral, 1 gain, 2 high-level gain/amplification), we observed highly significant (p < 1 ×10^−16^) expression-copy-number associations for canonical copy-number-driven oncogenes (*ERBB2*, *CCND1*, *FGFR1*, *MDM2*) and tumor suppressor genes (*MAP2K4*, *MTAP*, *PPP2R1A*), with Spearman ρ at range of 0.30 to 0.65 (Table S8). We then tested whether the prevalence of expression outliers differed between H/L and NHW in INSPIRE, focusing on 1185 genes in 93 gain regions and 1907 genes in the 122 loss regions with Spearman correlation between expression and copy number greater than 0.30 and found no genes met FDR < 0.05 (Table S8).

### Associations between germline variants with somatic mutations and COSMIC mutation signatures

Forty-five of 824 patients in INSPIRE carried pathogenic germline variants including 14 in *BRCA1*, 20 in *BRCA2*, 1 in both *BRCA1* and *BRCA2*, and 10 in *PALB2* (Table S9A). The most significant association with germline *BRCA1, BRCA2,* and *PALB2* variants was observed for COSMIC mutation signature SBS3 (p = 2.66 ×10^−19^ in multivariable regression analysis) with approximately five-fold higher proportion of the signature in the pathogenic variant group than in the non-pathogenic variant group (0.559 versus 0.109). Somatic mutations in *PIK3CA* were significantly less common in tumors with pathogenic *BRCA1, BRCA2,* and *PALB2* variants (p = 0.009 fisher exact test) and remained so after adjustment for PAM50 subtype (p = 0.04 multivariable logistic regression; Table S9B).

APOBEC3 is a gene family of mRNA editing enzymes highly expressed in cells of the human innate immune system. The germline *APOBEC3A/B* deletion creates an isoform of the APOBEC3A protein and deletes the APOBEC3B protein (Table S10A). We observed that this deletion was 2.58-fold more common in H/L than in NHW women (Fisher exact p = 9×10^−15^) (Table S10B) and was more common among H/L women with high IA ancestry (p = 3.09 ×10^−7^, Spearman correlation test) (Figure 4A). The frequencies of wildtype, heterozygous, and homozygous carriers of the deletion were 250 (57.21%), 150 (34.40%), and 36 (8.26%) in H/L and 317 (81.70%), 65 (16.75%), and 6 (1.55%) in NHW. The *APOBEC3A/B* deletion was associated with APOBEC related mutational signatures SBS2 (Figure 4B) and SBS13 (Figure 4C), consistent with previous reports by us and others^30,35^. We tested the association of the germline *APOBEC3A/B* deletion with somatic events, restricting the analysis to the 720 tumor samples from H/L (398 H/L from INSPIRE study and 322 H/L from CLBCS). Somatic mutations in *PIK3CA* were more common in patients with the *APOBEC3A/B* deletion (p = 0.003 in both the Cochran-Armitage trend test and multivariable logistic regression) (Table S10D). Among five *PIK3CA* mutations known to arise predominantly through APOBEC cytidine deamination (E365K, E453K, E542K, E545K, E726K), there was a highly significant increasing rate of mutation frequency across 0, 1, and 2 copies of *APOBEC3A/B* deletion (p = 0.00002), but there was no association with H1047R/L/Q/Y mutations (Figure 4D).

**Figure 4.**
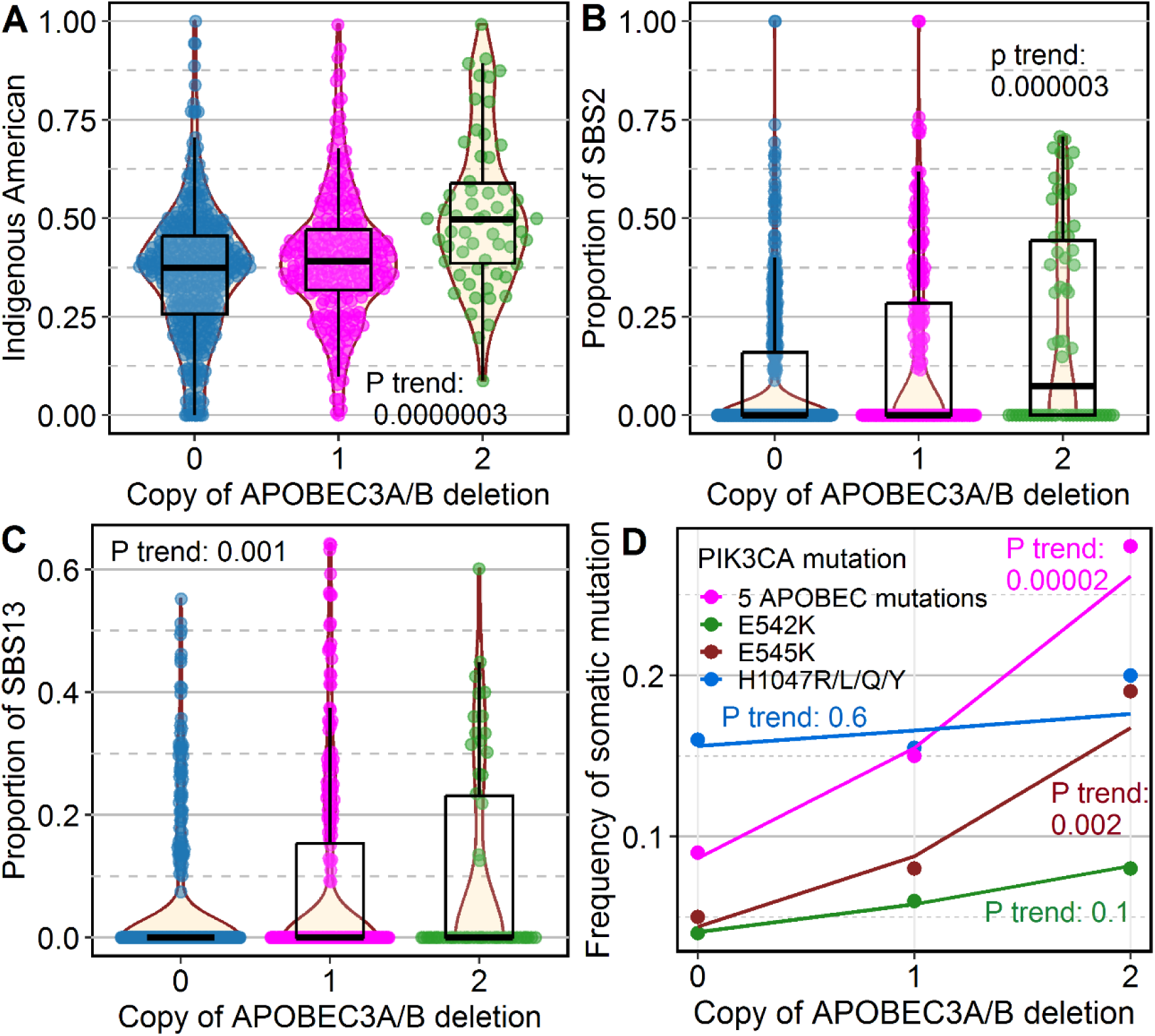
Associations of germline *APOBEC3A/B* deletion with: (A) Indigenous American (IA) ancestry. (B) COSMIC mutation signatures SBS2 and (C) SBS13, and (D) somatic mutations in *PIK3CA* in 720 Latinas (398 Latina samples from INSPIRE study and 322 Latina samples from CLBCS). In panels A-C, p values were generated by the Spearman correlation test in which *APOBEC3A/B* was coded as numeric number of 0, 1, and 2 copies of the deletion. In panel D, the five APOBEC-related mutations in PIK3CA are E365K, E453K, E542K, E545K and E726K; p values of association were generated by the Cochran-Armitage trend test.

### Ecotype immune signatures across tumor subtypes

To optimize evaluation of the TME from bulk transcriptomics, we used the Ecotyper program (https://ecotyper.stanford.edu/carcinoma/) which utilizes CIBERSORTx^36^ to calculate cell fractions based on RNAseq single-cell reference data, and then identifies 69 transcriptional cell states and their co-association patterns to discriminate 10 ecotypes (multicellular communities) ^37^. We used Ecotyper to identify TME states for the 1081 tumor samples with RNAseq data (759 from INSPIRE study and 322 from CLBCS, Table 1). Each tumor sample can have multiple ecotypes, with the dominant ecotype defined as the ecotype with the maximum abundance across the 10 ecotypes in a sample (Figure S2A). Across samples, ecotypes CE1, 2, 6, 8, 9, and 10 were common, whereas ecotypes CE3, 4, 5, and 7 had low frequencies; distributions of the ecotypes were significantly different among the tumors from the INSPIRE H/L, INSPIRE NHW, and CLBCS women (Figure S2B). The most common ecotypes in tumors were CE8 from both the INSPIRE H/L and CLBCS women, whereas the most common ecotype in tumors from INSPIRE NHW women was CE6. In analysis of the combination of 720 tumors from H/L women in the INSPIRE study and CLBCS, the distribution of dominant CE showed highly significant (P < 0.0001) differences in terms of tumor stage (Figure 5A), PAM50 subtypes (Figure 5B), and HER2 status (Figure 5C) (Table S11). The most common ecotype in the basal subtype was CE2 which was rare in other subtypes. CE6 was the predominant ecotype in the normal subtype and also was common in Luminal A tumors. CE8 was the most common in Luminal B and second most common in Luminal A. The HER2-enriched subtype had a mixture of different ecotypes and was notable for having a higher prevalence of CE10 than other ecotypes. CE2 high tumors are enriched in basal-like epithelial cells and associated with worse patient outcomes, CE6 high tumors are enriched for normal tissue, CE8 high tumors are associated with moderately favorable patient outcomes, CE9 and CE10 high tumors are pro-inflammatory and have been associated with longer survival in prior studies ^37^.

**Figure 5.**
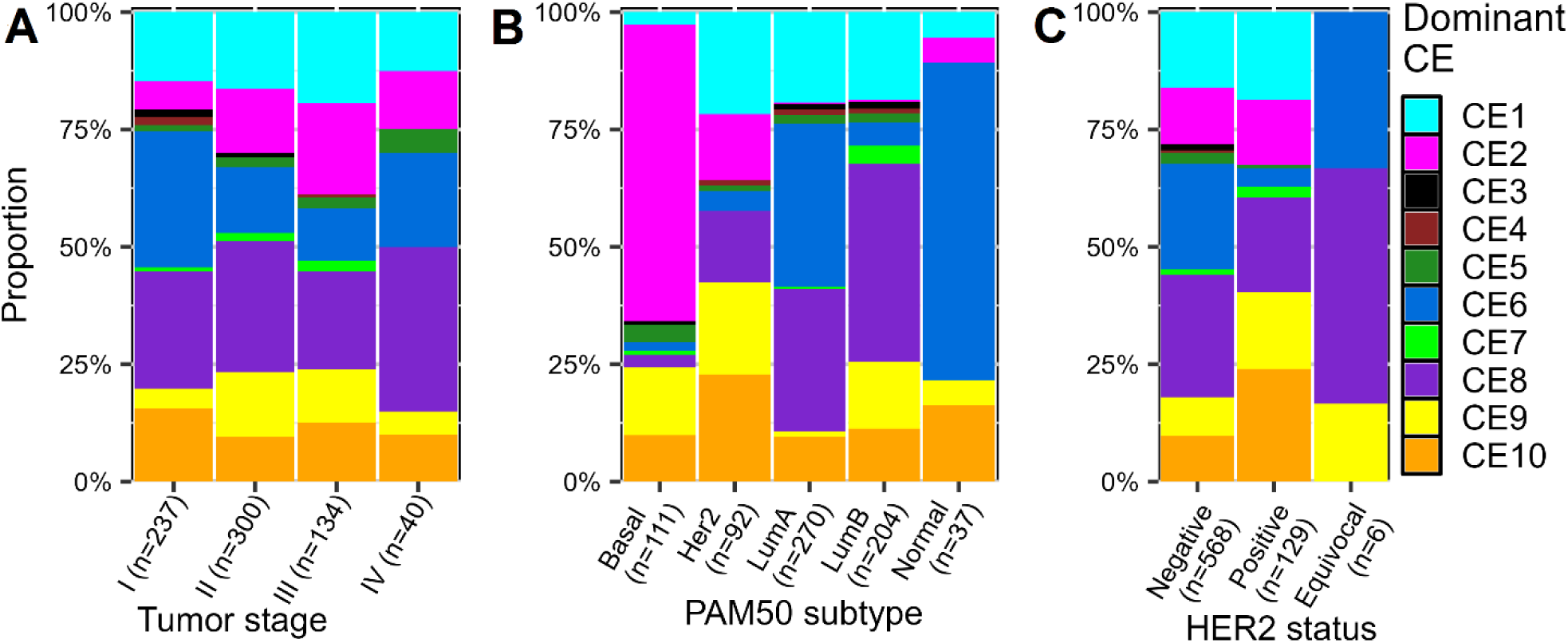
Distribution of dominant ecotypes by: (A) tumor stage; (B) PAM50 subtype; (C) IHC HER2 status in 720 Hispanic/Latinas from INSPIRE study and CLBCS. A dominant ecotype in a sample is the ecotype with the maximum abundance across the 10 ecotypes in a sample. Numbers within each subgroup are provided in Supplemental Table S11.

### Immune ecotypes, ancestry and recurrence

We investigated the association of the CE ecotypes with breast cancer recurrence using multivariable logistic or Cox regression analyses in the INSPIRE, CLBCS, and TCGA datasets. Associations of CE10 and the CE10 related cell states and marker genes with breast cancer recurrence risk were all significant (P < 0.05) in the CLBCS dataset (Figure 6B) and significant or marginally significant in the INSPIRE dataset (Figure 6A) and in the TCGA dataset (Figure S4C). Similar to CE9, levels of CE10 and the CE10-related cell states and marker genes were significantly higher in tumors from H/L compared to NHW women (Figure 6C). Among tumors from H/L women, CE10 and all its associated markers were significantly correlated with IA ancestry in analyses that combined INSPIRE and CLBCS (Figure 6D). CE10 and its associated markers were highly significantly (p < 0.0001) increased only in the HER2-enriched subtype (Table S12).

**Figure 6.**
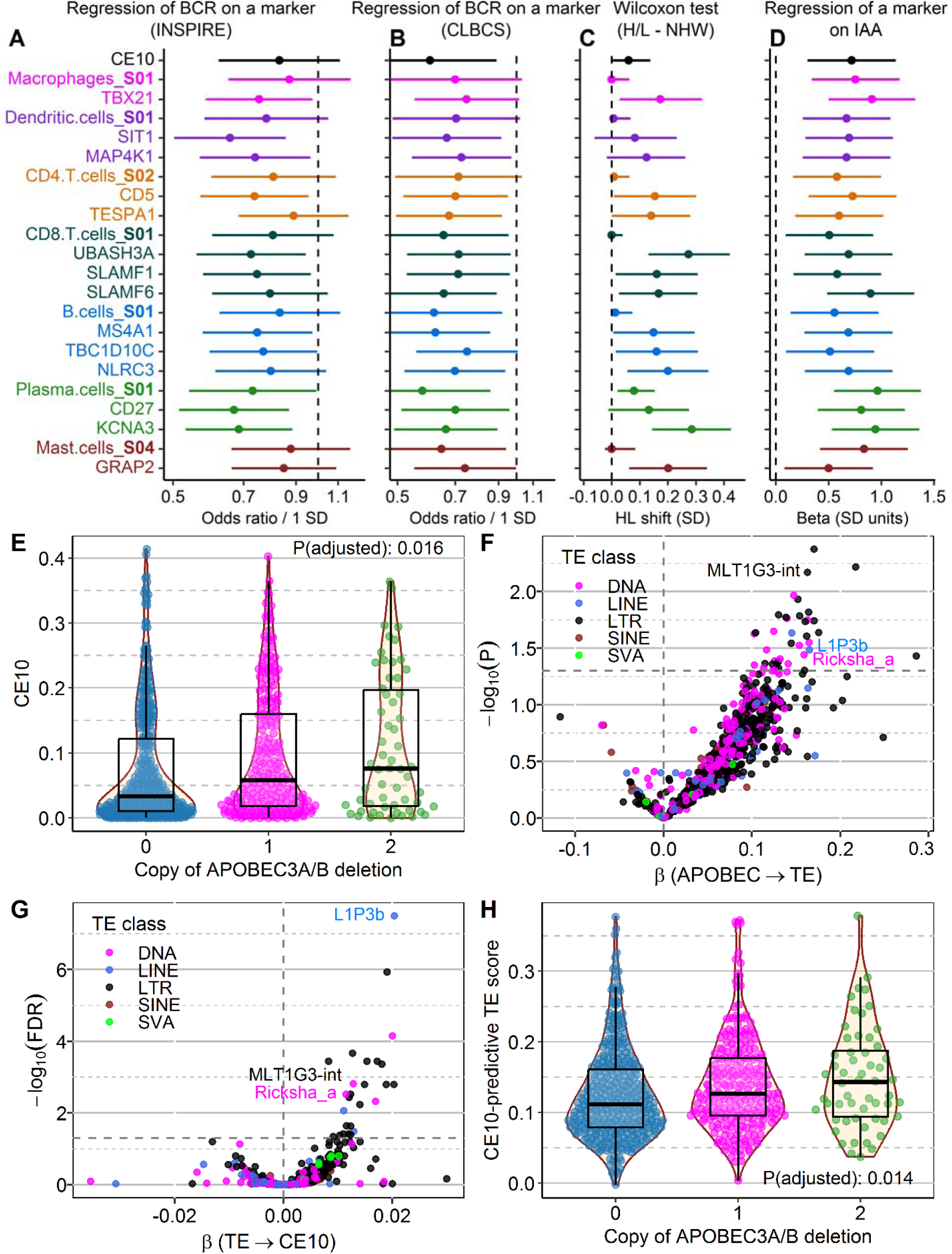
CE10 immune ecotype is: **(A, B)** prognostic for breast cancer recurrence (BCR) across cohorts, **(C)** enriched in Hispanic/Latinas (H/L) compared to non-Hispanic Whites (NHW), and **(D)** particularly in those of high Indigenous American ancestry (IAA), (**E**) significantly associated with *APOBEC3A/B* deletion and (**F**-**H**) transposable elements (TE). **A-D** are forest plots showing associations for CE10, the seven CE10-related cell states (S) and CE10-related genes for each cell state using point estimate with 95% confidence interval (horizontal bar) and vertical reference dash line. (**A,B**) Multivariable logistic regression demonstrated that CE10 and CE10-related genes and cell states are consistently associated with improved outcomes in **(A)** 759 samples from INSPIRE and **(B)** 312 H/L samples from CLBCS. For the logistic regression models, the x axis is the odds ratio (OR) per 1 standard deviation (SD) increase with 95% Wald CIs on a log scale (reference line OR = 1.0). (**C**) Wilcoxon test comparing H/L versus NHW women in INSPIRE. CE10 and CE10-related immune markers are significantly elevated in H/L tumors with the effect size estimated by the Hodges–Lehmann (HL) location shift in SD units on linear scale. (**D**) positive association of CE10-related immune markers with Indigenous American ancestry (IAA) in multiple regression with beta change for IAA on linear scale of SD (reference line of zero). For Panels A and B, multiple covariates include age at diagnosis, tumor stage, and PAM50 subtypes, as well as ethnicity in Panel A. CE10 **(E)** and CE10-predictive TE score **(H)** increase stepwise with the copy of APOBEC3A/B germline deletion; The TE score in H was derived from a LASSO model including 115 selected intergenic TEs, computed as a weighted linear combination of their expression levels; P values in panels E and H were obtained from multivariable linear regression models with CE10 (E) or TE score (G) as the outcome and APOBEC3A/B deletion as the predictor, adjusting for covariates. Volcano plots in F and G show associations between APOBEC3A/B deletion and TE expression from covariate-adjusted linear models (TE ∼ APOBEC3A/B deletion + covariates) (**F**) and associations between intergenic transposable element (TE) expression and CE10 (CE10 ∼ TE + covariates) (**G**); the x-axis in F and G show the regression coefficient (β) for APOBEC3A/B deletion and TE, respeactively; the y-axis shows -log□□(P value) in F and -log□□(FDR) in G. The dashed horizontal line indicates P value of 0.05 in F and FDR of 0.05 in G; three TEs labelled in both F and G shared P < 0.05 in F and FDR < 0.05 in G; In panels (F) and (G), points are colored by TE class (DNA, LINE, LTR, SINE, and SVA). Covariates in D-H are tumor purity, tumor mutation burden, IAA, and PAM50 subtype; samples in D-H are 720 H/L tumor samples (398 from INSPIRE and 322 from CLBCS).

High abundance of CE9 was significantly associated with lower risk of breast cancer recurrence among patients in the INSPIRE study (Figure 7A, Figure S3C), CLBCS (Figure 7B, Figure S3D) and TCGA (Figure S4A, Figure S5B). In addition, we analyzed the association of the cell states (S) and marker genes that defined the ecotypes and breast cancer recurrence. Within ecotypes, we ranked the top marker genes based on Spearman correlation (rho > 0.6) of a marker gene expression with corresponding ecotype abundance and then the consistency of association of marker genes with breast cancer recurrence in the three datasets (INSPIRE, CLBCS, and TCGA) (Table S12). CE9 and its associated cell states and marker genes had higher abundance or expression level in tumors from H/L women than in tumors from NHW women in both the INSPIRE study (Figure 7C, Figure S3A) and TCGA (Figure S4B, Figure S5A).

**Figure 7.**
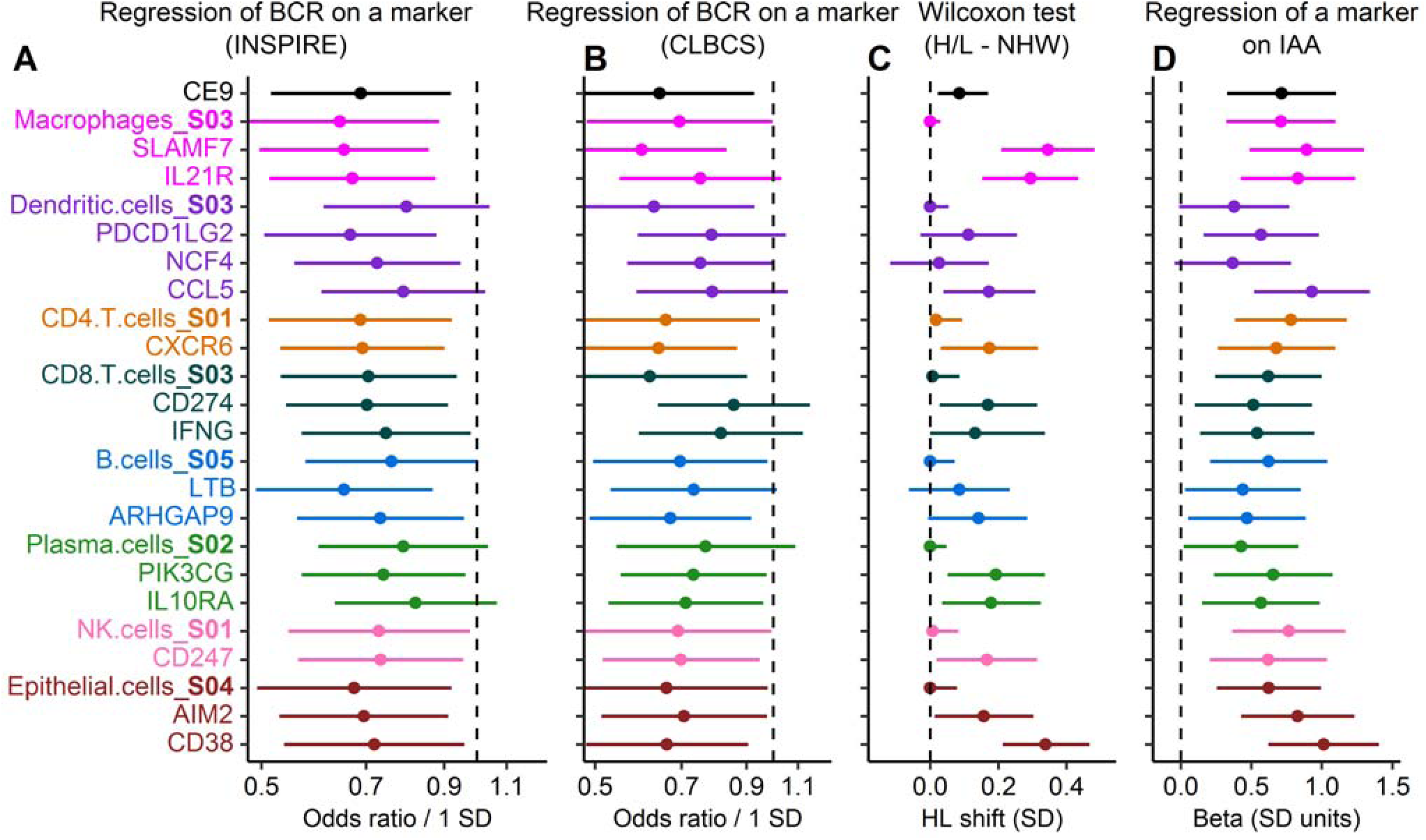
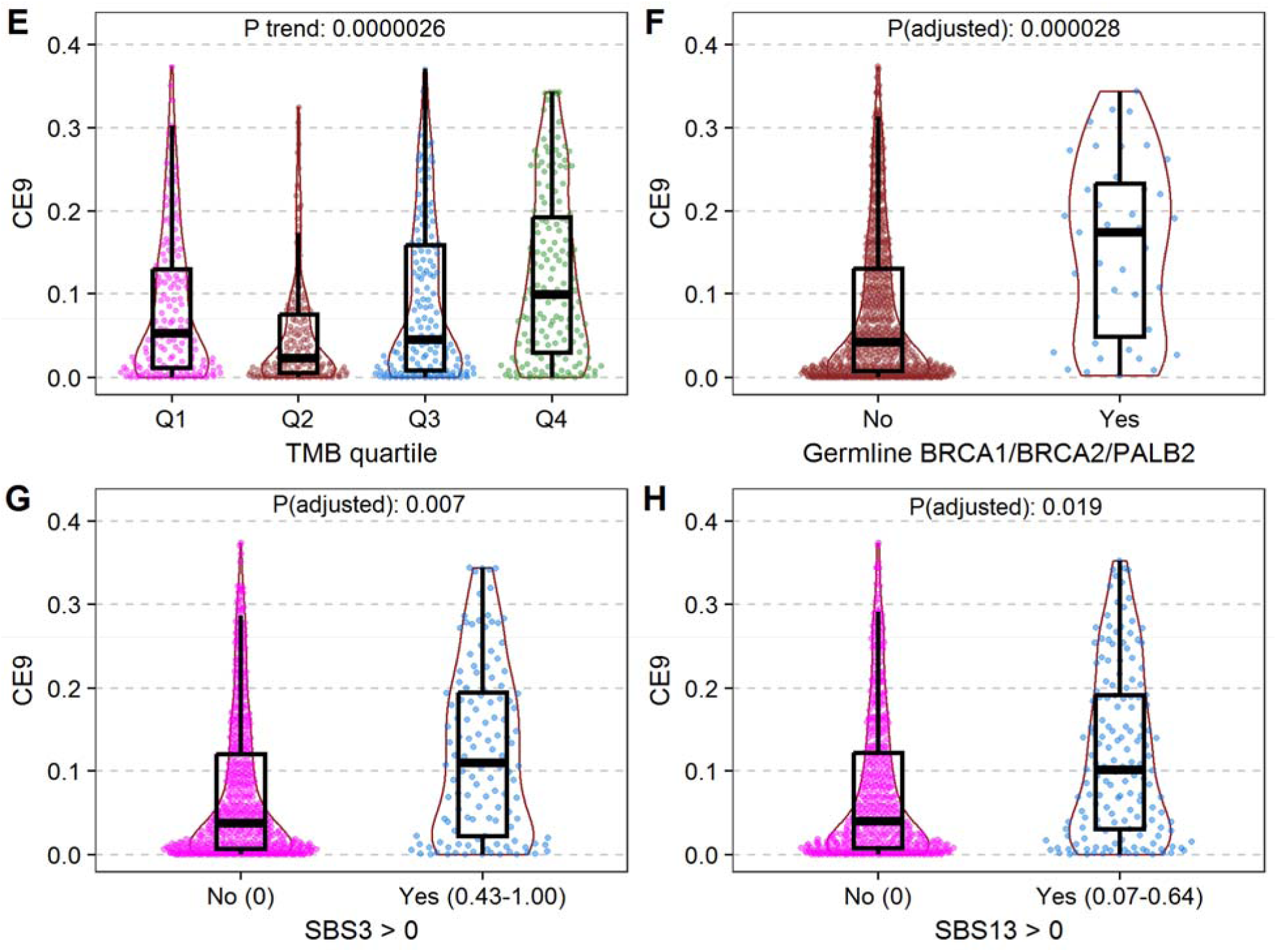
CE9 immune ecotype is: **(A, B)** prognostic for breast cancer recurrence (BCR) across cohorts, **(C)** enriched in Hispanic/Latinas (H/L) compared to non-Hispanic Whites (NHW), and **(D)** particularly in those of high Indigenous American ancestry (IAA), and significantly associated with **(E)** tumor mutation burden (TMB), **(F)** *BRCA1*, *BRCA2*, and *PALB2* pathogenic variants, **(G)** homologous recombination deficiency (HRD) COSMIC signature SBS3, and **(H)** APOBEC COSMIC signature SBS13. **A-D** are forest plots showing associations for CE9 ecotype, the eight CE9-related cell states (S) and CE9-related marker genes for each cell state using point estimate with 95% confidence interval (horizontal bar) and vertical reference dash line. (**A,B**) Multivariable logistic regression demonstrated that CE9 and CE9-related genes and cell states are consistently associated with improved outcomes in **(A)** 759 samples from INSPIRE and **(B)** 322 H/L samples from CLBCS. For the logistic regression models, the x axis is the odds ratio (OR) per 1 standard deviation (SD) increase with 95% Wald CIs on a log scale (reference line OR = 1.0). (**C**) Wilcoxon test comparing H/L versus NHW women in INSPIRE. CE9 and CE9-related immune markers are significantly elevated in H/L tumors with the effect size estimated by the Hodges-Lehmann (HL) location shift in SD units on linear scale. (**D**) for 720 H/L samples (398 from INSPIRE and 322 from CLBCS), positive association of CE9-related immune markers with Indigenous American ancestry (IAA) in multiple regression with beta change for IAA on linear scale of SD (reference line of zero). For panels **A, B** and **D**, multiple covariates include age at diagnosis, tumor stage, and PAM50 subtypes, as well as ethnicity in panel A. (**E**) CE9 significantly increased with tumor mutation burden (TMB) quartiles. (**F)** Association of CE9 with germline *BRCA1/BRCA2/PALB2* pathogenic variant status. Mutation carriers exhibited significantly higher CE9 levels. (**G**-**H)** Samples with detectable SBS3 or SBS13 showed elevated CE9. P-value in panel E was from regression of CE9 on numeric TMB quartile variable. P values in panels F to H were generated by regression of CE9 on: **(F)** mutation status, **(G)** SBS3, **(H)** SBS13, adjusting for tumor purity, TMB, IA ancestry and PAM50 subtype, respectively.

Among all tumors from H/L women in the combined INSPIRE and CLBCS datasets, CE9 and its cell states and marker gene levels were significantly associated with higher IA ancestry after adjusting for age at breast cancer diagnosis, tumor stage and PAM50 subtype (Figure 7D, Figure S3B, Table S12). In multiple regression analyses of CE9, CE9-associated cell states, and CE9 marker genes on age at breast cancer diagnosis, tumor stage and PAM50 subtype, high abundance of the CE9 markers were highly significantly (p < 0.00001) enriched in samples with non-Luminal A subtypes (Luminal B, Basal and HER2-enriched subtypes) in the INSPIRE, CLBCS, and TCGA datasets (Table S12). Some CE9-associated marker genes such as SLAMF7 and CD38 consistently demonstrated stronger associations than CE9 (Figure 7A-D, Figure S4A-D, and Table S12) with breast cancer recurrence risk (Figure 7A-B), ethnicity (Figure 7C), and IA ancestry (Figure 7D) in datasets from INSPIRE, CLBCS and TCGA (Figure S6 and S7).

### Drivers of immune ecotypes

The *APOBEC3A/B* deletion was significantly associated with higher CE10 (Figure 6E) (p value = 1.74 × 10^−4^, FDR < 0.001) (Table S10C). We also performed multivariable linear regression of CE10 ecotype on *APOBEC3A/B* deletion and the association remained significant (p = 0.016 after adjusting for IA ancestry, age at diagnosis, tumor stage, PAM50 subtype, and TMB (Table S10E). The association of CE10 with IA ancestry was attenuated by adjusting for *APOBEC3A/B* deletion in the model, consistent with the higher frequency of the deletion in IA ancestry and its strong association with CE10 (Table S10E). We evaluated the effect of APOBEC signatures SBS2 and SBS13 on CE10 to determine if the association between *APOBEC3A/B* deletion and CE10 may be due to its effect on mutational patterns. In analyses that included all *APOBEC3A/B* deletion genotypes, there was a nominally significant association with SBS2 (Figure S10A) but not with SBS13 (Figure S10C). There was no association of CE10 with SBS2 and SBS13 among women with no *APOBEC3A/B* deletions (Figure S10B and S10D). In the subset of the tumors with no SBS2 or SBS13, *APOBEC3A/B* del still had a trend towards association with CE10 (Figure S10E). Therefore, the association did not seem to be driven by the mutational signatures associated with the *APOBEC3A/B* deletion.

Since APOBEC enzymes suppress viruses, including transposable elements (TEs) ^38,39^, and since reactivated TEs have been associated with increased immune response ^40^, we hypothesized that the *APOBEC3A/B* genotype association with immune ecotype CE10 subtype was based on its loss of suppression of TEs. First, we tested the association between *APOBEC3A/B* deletion and TE expression. The majority of TEs exhibited positive associations with *APOBEC3A/B* deletion in models regressing individual TE expression on *APOBEC3A/B* deletion, adjusting for the same covariates (Table S13A; Figure 6F). Although no individual TE was associated with *APOBEC3A/B* deletion at an FDR <0.05, all 40 of the TEs that were nominally associated (p < 0.05) had higher expression with the deletion (Table S13A). Gene set enrichment analysis revealed a significant enrichment of TEs classified as long terminal repeats (LTRs) among TEs positively associated with *APOBEC3A/B* deletion (adjusted *p* = 0.0004; normalized enrichment score = 1.46, Table S13B). The vast majority of LTRs (550 of 578; 95.2%) were members of one of the families of endogenous retroviruses (ERVs). In addition, DNA TEs were significantly enriched for associated with *APOBEC3A/B* deletion (adjusted p = 0.008; normalized enrichment score = 1.35, Table S13B).

Next, we tested for association between the TEs and CE10. A total of 26 out of 1,052 transposable elements (TEs) were significantly associated with CE10 (FDR < 0.05), and all exhibited positive associations (i.e., increased expression with higher CE10), in models regressing CE10 on individual TE expression while adjusting for tumor purity, IA ancestry, TMB, and PAM50 subtype (Table S13A; Figure 6G) using H/L samples. To validate these findings, we examined the association between CE10 and TEs in TCGA samples and found that 14 of the 26 TEs replicated at FDR < 0.05, with an additional two reaching nominal significance (P < 0.05). Importantly, all 16 TEs showed positive associations with CE10 in TCGA, consistent with the direction observed in our dataset, supporting the robustness and reproducibility of these associations (Table S13A).

From enrichment analysis in our data, we identified LTRs as significantly enriched (adjusted p = 4.0×10□□, normalized enrichment score = 1.69, Table S13B). Notably, *L1P3B*, the TE most strongly associated with CE10 (FDR < 2.6×10^−14^) was also positively associated with the *APOBEC3A/B* deletion (p = 0.033). Two other TEs, *MLT1G3-int* and *Ricksha_a*, were associated with CE10 at FDR < 0.05 and with the *APOBEC3A/B* deletion at p < 0.05. To capture the joint contribution of TE expression on CE10, we applied LASSO regression to model CE10 as a function of all 1,052 TEs, adjusting for the same covariates. We identified 115 TEs that together explained approximately 77% of the variance in CE10. Then we computed a CE10-predictive TE score as a weighted sum of the expression levels of these 115 TEs, with weights given by the coefficients estimated from the LASSO model. The CE10-predictive TE score was significantly and positively associated with the *APOBEC3A/B* deletion (adjusted p *=* 0.014; Figure 6H). Mediation analysis demonstrated that the TE-derived composite score significantly mediated the association between the *APOBEC3A/B* deletion and CE10 (average causal mediation effect, p = 0.021), accounting for approximately 75% of the total effect, while the direct effect was not statistically significant (p = 0.168) (Table S13C). Collectively, these findings suggest that the *APOBEC3A/B* deletion may shape the CE10 immune ecotype through coordinated upregulation of transposable element expression.

CE9 increased progressively across quartiles of tumor mutation burden (TMB) (Figure 7E). Carriers of germline pathogenic variants in *BRCA1, BRCA2,* and *PALB2* were significantly more likely to have ecotype CE9 and its seven co-occurring cell states (p values ranged from 0.000003 to 0.001) with an approximately two-fold higher proportion of CE9 and cell states in the mutation group than the non-mutation group (Figure 7F, Table S9C). In addition, SBS3, which is considered a marker for HRD, was predictive of CE9, even after adjustment for TMB (Figure 7G). This association was also consistent among women who did not carry a pathogenic variant in *BRCA1, BRCA2,* or *PALB2* (Figure S8). CE9 was significantly elevated in tumors with the APOBEC-related SBS13 signature even after adjustment for TMB (Figure 7H). CE9 was not associated with SBS2 (Figure S9) or with *APOBEC3A/B* deletion (Table S12).

## DISCUSSION

We analyzed data from two large cohorts of women with breast cancer including the INSPIRE cohort with 436 H/L women and 388 NHW women and the CBCLS with 312 H/L women. We systematically evaluated somatic mutations, mutational signatures, and tumor immune ecotypes inferred from transcriptomics and comparisons between H/L and NHW women and by IA ancestry.

The majority of somatically mutated genes and copy number aberrations were consistent among H/L and NHW women. Our somatic mutational analyses identified inactivating mutations in *CTCF* as more common in H/L women than NHW women, although this did not reach statistical significance after correction for multiple hypothesis testing. CTCF is a zinc finger nuclear protein that acts as a transcription factor, chromatin remodeler, and insulator protein ^41–43^. Mutations in the 11 zinc fingers affect both direct and indirect site-specific effects on binding accessibility ^44^. The mutations we observed included stop codons and frameshift mutations with the vast majority in zinc finger domains. As in a prior study ^45^, we found that *CTCF* mutations were more likely in tumors with Luminal A and B subtypes. For *TP53*, the higher rate of mutations in the H/L samples compared to NHW in the INSPIRE dataset was explained by different proportions of PAM50 subtypes between the two ethnic groups and was not associated with IA ancestry. This highlights the need to incorporate covariates in analyses that may explain differences between populations. Most of the significantly mutated somatic genes in breast tumors were not different by self-reported ethnicity and/or not correlated with ancestry in our study. This suggests that despite the known environmental and genetic differences in exposures, the strongest mutational influences both in terms of individual genes and mutational patterns in tumors remain relatively consistent in H/L and NHW women.

Despite the similarity in somatic mutations and copy number aberrations among H/L women and NHW, we saw significant differences in between these groups in tumor immune ecotypes. Both CE9 and CE10 immune ecotypes were more common in H/L women compared to NHW women, and among the H/L women, those with the highest IA ancestry had higher CE9 and CE10 ecotypes. The association between IA and CE10 is partially explained by the *APOBEC3A/B* deletion, demonstrating that germline genetic variation may account for some differences in TME across populations of different ancestries. These results also demonstrate the advantage of inclusion of different ancestry populations in tumor profiling since the common frequency of the *APOBEC3A/B* deletion in H/L enhanced power to detect the association with CE10. The association between IA and CE9 is not explained by germline variants and/or somatic mutational signatures that we identified as predictors of CE9 in this population and suggest other genetic and/or environmental factors may underlie this association. Since CE9 is a strong predictor of response to immune checkpoint inhibitors, identifying and testing for these factors may identify women who are more likely to respond to immune checkpoint blockade.

Immune ecotypes in the TME are strong predictors of cancer survival and response to both chemotherapy and immune checkpoint inhibitors ^37,46–48^. In previous studies of lung cancer, melanoma, urothelial cancer, and ovarian cancers, tumors with CE9 and/or CE10 ecotypes had more favorable outcomes in ^37,49–51^. Both CE9 and CE10 and their co-occurring cell states are immune signatures enriched in CD4, CD8 T cells, macrophages, and dendritic cells in the TME and characterize proinflammatory classes of tumor phenotypes with favorable response to immune therapy ^37^. Consistent with prior studies in other cancers, we found that high abundance of CE9, CE9-associated cell states, and marker genes were significantly associated with lower risk of breast cancer recurrence among patients in the INSPIRE study, CLBCS, and TCGA datasets. For CE10, related cell states and marker genes were significantly associated with lower risk of breast cancer recurrence risk (P < 0.05) among patients in the CLBCS, and significant or marginally significant in the INSPIRE and TCGA. CE9 was highly enriched in both HER2-enriched and Basal subtypes, and CE10 was more enriched only in the HER2-enriched subtype in the INSPIRE, CLBCS, and TCGA data.

Our analyses revealed novel drivers of these tumor immune ecotypes. We identified a strong association between the *APOBEC3A/B* deletion and CE10. Prior studies have identified an association between the APOBEC mutational signature, germline deletion, and immune cell infiltration^33–37^ and hypothesized that it may be via the effect on mutational processes. The *APOBEC3A/B* deletion strongly predicted the APOBEC cosmic signatures SBS2 and SBS13. However, we found no association between the APOBEC mutational signatures, SBS2 and SBS13, and CE10 in patients not carrying the *APOBEC3A/B* deletion, suggesting that the mutational signatures were not the etiology.

APOBEC enzymes are cytosine deaminases that convert cytosine to uracil and function to suppress replication of viruses including ERVs in human TEs ^38,39^. TEs are frequently overexpressed in tumors and are associated with a tumor immune response (PMID: 31745090). In our analyses, the *APOBEC3A/B* deletion was associated with increased expression of many TEs, with approximately 50% being ERVs. Furthermore, a subset of ∼20 ERVs were very strongly associated with CE10, explaining ∼70% of the variance of this immune ecotype. The TEs which were the strongest predictors of CE10 were preferentially associated with the *APOBEC3A/B* deletion. These results suggest that CE10 is driven, in large part, by TEs including ERVs and that the *APOBEC3A/B* deletion is associated with CE10 due to derepression of TEs. However, the TEs were much stronger predictors of CE10 than *APOBEC3A/B* deletion, suggesting that additional factors may drive the association between the TEs and CE10.

CE10 ecotype is a pro-inflammatory tumor immune subtype, but in contrast to CE9, another pro-inflammatory subtype, it also is characterized by T cell markers of naive and central memory cells and with pro-inflammatory monocytes, cDC1 dendritic cells, and naive/resting B cells. The association with TEs suggests that this subtype of immune response may be more specific to retroviral expression.

We found that *BRCA1*, *BRCA2*, and *PALB2* pathogenic variants and SBS3 (the mutational signature of HRD) were associated with CE9. The association with SBS3 was consistent regardless of whether *BRCA1*, *BRCA2*, and *PALB2* pathogenic variants were present, suggesting that the mutational process itself in an important determinant of CE9. This result demonstrates that, in addition to total number of mutations, the type of mutations and underlying mutational processes influence the CE9 ecotype. Tumors with HRD also have substantial defects in chromosomal copy number^18^ which may contribute to the tumor immune landscape.

Several of the markers underlying CE9 that are strongly associated with IA ancestry and survival may be candidates for improving treatment response. The gene markers most strongly correlated with CE9 were *SLAMF7* and *CD38*. High expression of SLAMF7 directs macrophages to engulf solid tumor cells ^52^ which enhances antigen presentation and may improve immunotherapy outcomes. CD38 expression in T cells has been linked to T-cell exhaustion and dysfunction, and blockade of CD38 has enhanced immune checkpoint inhibition in pre-clinical studies ^53^. High expression of CD38 is associated with better responses to treatment using ICI ^54^. In addition, high expression of the other six CE9-related immune TME markers genes, CXCR6, IFNG, CD247, LTB, CCR5, and IL10RA, also showed prognostic and therapeutic potential for different type of cancers ^55–59^.

Our study represents the largest analysis of breast tumor genomics and transcriptomics among H/L women living in California, predominantly of European and IA ancestry. We focused on H/L women and compared to NHW, all from the INSPIRE dataset and added H/L women from the CBCSL to improve power in that population. There were some limitations. Our population was hospital-based and had low levels of African admixture. Future studies in other H/L populations with mixed African ancestry may identify different factors associated with the genetic and/or environmental factors that contribute to tumor landscapes in those populations. Our analyses of the immune TME were based on bulk transcriptomics which may not detect the level of detail available from surveys of individuals cells in tumors. In addition, bulk transcriptomics may miss crucial differences in the spatial distribution of cells which may affect tumor outcomes. Lastly, because our genomic analyses did not include array data or whole genome sequencing, we were limited in our ability to make conclusions about copy number and non-coding mutation differences.

In summary, we analyzed 748 H/L tumor and matched normal exomes and evaluated tumor transcriptomics from City of Hope patients. Overall, the somatic profiles in breast tumors from H/L women were similar to NHW women in our study. However, we did observe some specific differences. We found that *CTCF*, a tumor suppressor and recurrently mutated gene in breast cancer, was more commonly mutated in H/L women after accounting for covariates. We also identified that the germline *APOBEC3A/B* deletion was significantly more common in H/L women than NHW women and was strongly associated with IA ancestry. Importantly, we identified that immune TMEs that are associated with better prognosis and response to ICI therapy were more common in H/L women compared to NHW women and that among H/L women, they correlated with IA ancestry. Our results represent an important advance in understanding of the tumor immune microenvironment, tying germline genetic variation, APOBEC activity, TE and tumor immune response.

## METHODS

### Participants

Breast cancer participants were patients treated at City of Hope (COH), consented, and enrolled in IRB-approved studies. Of the participants, 824 were from the Implementing Next-generation Sequencing for Precision Intervention and Risk Evaluation (INSPIRE) study, a prospective cohort study of cancer patients where clinical genetic testing, tumor/normal whole exome sequencing, and RNA sequencing was performed. Within INSPIRE, we selected a subset of 436 breast cancer patients who self-identified as H/L and 388 who self-identified as NHW. The second study is the California Latina Breast Cancer Study (CLBCS) and includes 312 self-identified H/L COH breast cancer patients. Inclusion criteria were: 1) self-identified as H/L and 2) their tumor tissue from surgery was available and the sample contained more than 40% tumor based on examination by a single breast pathologist (D. Schmolze). For the CLBCS samples, we previously had reported on 140 of them ^30^. For all participants, clinical data were abstracted from medical records including age at diagnosis, tumor stage, histological estrogen receptor (ER), progesterone receptor (PR), and human epidermal growth factor (HER2) status, and breast-cancer recurrence status. Three of the 824 breast-cancer patients from INSPIRE and 10 of 312 breast-cancer patients from the CLBCS study had two primary breast cancers with tissue available for study.

### Whole exome DNA sequencing (WES) and whole-genome RNA sequencing

For the INSPIRE study, tumor DNA/RNA were extracted from FFPE tissue using Qiagen AllPrep DNA/RNA FFPE Kit after a pathologist evaluated the tumor contents on the tissue block. Tumor-matched normal DNA was extracted from peripheral blood or saliva for WES. The minimum input was 50 ng with the A260/280 ratio range of 1.8 to 2.0 for DNA sequencing and 25 ng with a ≥ 20% DV200 value for RNA sequencing. DNA libraries for WES were prepared using the KAPA HyperPrep library kit (Roche). RNA libraries were prepared using the KAPA RNA HyperPrep with Riboerase kit (Roche) for Total RNA. For the INSPIRE study, DNA and RNA sequencing were generated using the GEM ExTra (**G**enomic **E**nabled **M**edicine **Ex**ome and **Tra**nscriptome) assay (Ashion/Exact Sciences) ^60^. Ashion/Exact Sciences conducted the WES with 400X average coverage for somatic variants (800X average coverage across 442 cancer genes and 300X over rest of genome) and 200X average coverage for germline variants. Total RNA sequencing reads were greater than 100 million per sample. Tumor/germline sequence data were aligned to the human genome (Build37). Germline variant calling from the BAM files was performed using GATK Haplotype Caller. Somatic mutations were called using the FreeBayes workflows^61^. All data are stored in the COH POSEIDON database. More detailed descriptions of WES for CLBCS and the INSPIRE study are available from previous publications by Ding et al and White et al, respectively ^30,60^.

### Genetic ancestry analysis using germline SNPs

We performed genetic ancestry estimation for each of the 824 patients from the INSPIRE study and the 312 patients from the CLBCS using the germline WES data. We downloaded the whole genome sequence (WGS) data for 929 HGDP (Human Genome Diversity Project) samples from 54 diverse human populations as reference data. A combined gvcf file was generated for patient samples by the GATK HaplotypeCaller (https://software.broadinstitute.org/gatk). We then extracted overlapping SNPs between the WES for patient samples and the WGS data for the HGDP reference samples and further refined the SNP list using the genotype call rates and pairwise linkage disequilibrium between SNPs with a total of 54,931 shared SNPs for analysis. We first used ADMIXTURE v1.3.0 to estimate underlying ancestral populations. The optimal number of ancestral components (K) for the reference samples was determined using five-fold cross-validation. The cross-validation error reached a minimum at K ≈ 8 - 9, indicating that the SNP genotype data support approximately eight to nine stable ancestral clusters in the reference population. We then used ADMIXTURE 1.3.0 to estimate genetic ancestry proportions by the best k value of eight on the combined data from reference and test samples. In addition, we used the principal components analysis, as implemented in PLINK 1.9 ^62^, as a complementary method to assess population structure.

### Germline *APOBEC3A/B* deletion calling

We assigned genotypes for the *APOBEC3A/B* deletion for all samples in the INSPIRE and CLBCS data based on the distribution of reads in the deletion region. The deletion causes loss of the 3’ end of *APOBEC3A* and loss of all but the 3’ end of *APOBEC3B*^63^. Genotypes of the germline deletion of *APOBEC3A/B* were assigned for samples from INSPIRE and CLBCS based on histogram distribution of reads ratio between the 30Kb deletion regions and its flanking regions in normal sample in the pileup file, which was the ratio of total number of reads in the deletion regions (3’UTR of *APOBEC 3A*, Exon 1 to exon 7 *of APOBEC 3B*) and total number of reads in the flanking non-deletion regions (exon 1 to exon 4 of *APOBEC 3A* and 3’UTR of *APOBEC 3B*) (Table S10A, Figure S11). Loss or reduction of gene expression of *APOBEC3B* confirmed loss of both or one copy, respectively. We previously published genotype calling for this deletion in 140 H/L ^30^based on genotype of rs12628403 which showed high linkage disequilibrium with the APOBEC deletion^64^. Genotype calling using distribution of sequence reads was consistent with previous calling using the SNP rs12628403.

### RNAseq data processing, PAM50 subtype inference, and Ecotype inference

RNAseq reads were aligned to hg19 genome assembly using Tophat2 (v2.0.8) with default settings. Gene-expression levels were counted by obtaining raw counts with HTSeq (v0.6.1p1) against Ensembl v86 annotation. The raw counts data were normalized using the trimmed mean of M values (TMM) method implemented in R package edgeR. Log2-transformation of normalized counts (log2FPKM) were used to assign PAM50 subtypes for each tumor sample based on the subgroup-specific gene centering method developed by Zhao, *et al.* Log2(FPKM) gene expression matrix with genes in rows and samples in columns served as input for the Carcinoma EcoTyper program (https://ecotyper.stanford.edu/carcinoma/) to identify and quantify proportion of pre-defined transcriptional cell states and cell ecotypes in each tumor sample.

### Somatic copy number analysis

Using the BAM file for each matched tumor-normal sample pair, we generated a pileup file summarizing the counts of reads with the reference (ref) allele, alternate (alt) allele, errors (neither ref nor alt), and deletions at each specific genomic position covered by the Exome baits. This pileup file was used for the FACETS program, implemented in R package FACETS version 0.6.1, to calculate somatic copy number alteration (SCNAs) for each tumor sample. The segmentation files generated by FACETS served as input files for GISTIC2.0 ^65^ on the GenePattern server (https://genepattern.broadinstitute.org/gp) to identify significant SCNAs using a q-value cutoff < 0.05. Gene-level somatic copy-number scores (states) were assigned using GISTIC2-thresholded values: −2 (deep loss), −1 (shallow loss), 0 (neutral), 1 (low-level gain), and 2 (high-level gain/amplification). Expression outliers were defined by robust Z-scores (Z > 2.0 for amplification; Z < −2.0 for deletion). Outliers were classified as copy-number–driven when gene expression showed a strong monotonic association with the GISTIC2-thresholded score (Spearman ρ > 0.30, P < 1×10□^1^□), using cutoffs derived from well-established CN-driven genes (*ERBB2*, *CCND1*, *FGFR1*, *MDM2*, *MAP2K4*, *MTAP*, *PPP2R1A*) ^2^. Fisher’s exact test was used to identify genes with frequency difference in expression outliers, driven by copy-number alterations, between 398 tumor samples from H/L and 361 NHW in INSPIRE study. Adjusted p value (FDR) was calculated by Benjamini-Hochberg method ^66^.

### TCGA breast cancer data

Somatic mutation data for TCGA breast cancer samples were obtained from the “maftools” R package. Raw counts of RNAseq data for TCGA breast cancer samples (including tumor and matched normal samples) were downloaded from the Genomic Data Commons (GDC) using the GDCRNATools R package ^67^. We downloaded the ecotype data for TCGA breast cancer samples and the list of core genes specific to breast cancer that define different cell states in each of 12 cell types (CD8.T.cells, CD4.T.cells, Fibroblast cells, NK cells, Plasma Cells, Epithelial cells, Dendritic cells, Mast cells, Monocytes and Macrophages cells, Polymorphonuclear cells (PMNs), B cells, Endothelial cells) from the EcoTyper study (https://ecotyper.stanford.edu/carcinoma/).We previously published genetic ancestry data for TCGA *BRCA* breast cancer samples ^30^.

### Demographic and clinical features of H/L and NHW participants

We tested association of self-reported ethnicity (H/L versus NHW) with age at diagnosis, ER status, PR status, HER2 status, ER/PR/HER2 triple negative status, tumor stage, PAM50 subtypes, and breast cancer recurrence. Association of age at diagnosis with self-reported ethnicity was tested using the Wilcoxon rank sum test and all the other associations between two categorical variables were tested by the Fisher’s exact test; For association of tumor stage with ethnicity, we further tested the association by adjusting age at diagnose in the multinomial logistic regression analysis with tumor stage as the outcome, and ethnicity plus age at breast cancer diagnosis as two independent predictors. PAM50 subtype, inferred based on RNAseq data, were available only for 759 of 824 patients; Breast cancer recurrence was evaluated only for patients with tumor stages I, II and III. In addition to using fisher exact test to check association between recurrence status and ethnicity, we further tested the association by adjusting age and tumor stage using binary logistic regression with the recurrence status as outcome and ethnicity, age at diagnosis, and tumor stage as predictors.

### Somatic mutations and somatic mutation signatures in H/L and NHW tumor samples

We identified somatic single nucleotide variants (SNV) and short insertion/deletion variants and further filtered by read depth ≥ 10, and reads for alternative allele ≥ 4, and the alternate allele ratio ≥ 0.1. We performed a somatic-mutation significance analysis using MutSigCV (version 1.3.5) on Genepattern (https://www.genepattern.org/modules/docs/MutSigCV). Genes with false discovery rate (FDR) q < 0.05 were considered to be significantly mutated genes. We counted the number of mutations and their corresponding mutation frequencies in the self-reported H/L and NHW groups and performed Fisher’s exact test to investigate if any gene was significantly more frequently mutated in either ethnic group. For genes with significant association in the Fisher exact test, we further performed multivariable logistic regression analysis to test the association by including five covariates (neoadjuvant treatment, age at diagnosis, proportion of IA ancestry, tumor stage, and PAM50 subtype). Furthermore, for each ethnic group, we tested whether somatic mutations in a gene were associated with IA genetic ancestry using logistic regression models in which mutation (Yes or No) was the outcome variable and numerical global ancestry proportion were the explanatory variables.

For each tumor sample with ≥ 10 somatic SNVs, we assigned COSMIC single-base-substitution (SBS) mutation signatures using the SigProfilerAssignment tool ^68^ for the 30 version-2.0 SBS mutation signatures. We tested differences in SBS mutation signatures between H/L and NHW using the Wilcoxon test.

### Ecotype signatures in tumor microenvironment (TME)

Log2(FPKM) gene expression matrix with genes in rows and samples in columns served as input to identify and quantify proportion of pre-defined 69 transcriptional cell states and 10 ecotypes in each tumor sample by the Carcinoma EcoTyper program (https://ecotyper.stanford.edu/carcinoma/). Cell states within a cell type in TME were defined by specific gene expression signatures with clusters in columns and a list of marker genes in rows; they were generated from deconvolving multi-sample bulk RNAseq data with insights from published single-cell data. Then a group of statistically co-occurring cell states across multiple cell types in the TME further formed into an ecosystem subtype or ecotype. We first tested association of abundance of each ecotype (numeric variable) with status of breast cancer recurrence (Yes/No) for INSPIRE samples adjusting for self-identified ethnicity, genetic ancestry, age at diagnosis, tumor stage and PAM50 subtypes using multivariable logistic regressions; the same logistic regressions were performed for CLBCS samples. For TCGA samples, we tested association of abundance of each ecotype (numeric variable and quartile variable) with overall survival and progression-free survival adjusting for ethnicity, age at diagnosis, tumor stage and PAM50 subtypes using multivariable Cox regression analysis. Then we examined the association of abundance of each ecotype with the four covariates listed above using multivariable linear regression analysis. Finally, if an Ecotype consistently showed significant associations with breast cancer outcomes in the INSPIRE, CLBCS and TCGA studies, we investigated molecular mechanisms of each ecotype. We performed the same two analyses described above for both the Ecotype-associated cell states and marker genes that define each cell state. We also tested ethnicity-specific differences in ecotypes/ and their marker genes using the Wilcoxon test for INSPIRE data and TCGA data. For the 398 H/L in INSPIRE and the 322 H/L in CLBCS, we tested the association of IA ancestry with ecotypes/cell states and their marker genes by linear regression of each marker on age, IA ancestry, tumor stage and PAM50.

### Association of pathogenic germline variants with somatic mutations, COSMIC SBS mutation signatures, and ecotype signatures

Pathogenic germline variants in *BRCA1*, *BRCA2*, and *PALB2* in the INSPIRE study were identified from clinical genetic testing of the Invitae Multi-Cancer panel. Using Fisher exact test and multivariable logistic regression analysis, which was restricted to genes with at least 5% somatic mutations, we tested the association of pathogenic *BRCA1/BRCA2/PALB2* pathogenic variants (Yes/No status) with somatic mutations (Yes/No status) in the 22 cancer driver genes identified by MutSigCV. For mutation signatures and ecotype signatures, we performed multivariable linear regression of continuous abundance of signatures on breast cancer status (Yes/No) adjusting for age at diagnosis, self-reported ethnicity, genetic ancestry, tumor stage, PAM50 subtype and TMB. TMB is defined as the number of non-silent somatic non-synonymous single-nucleotide variants (SNVs) and small insertions/deletions normalized by the bait capture size.

The Cochran-Armitage trend test was used to test the association of the number of *APOBEC3A/B* deletion allele (0,1,2) with categorical variables including somatic mutations (Yes/No status) in cancer driver genes, ER/PR/HER2 status, and PAM50 Her2 subtype. For the genes with significant association at P < 0.05, we further performed multivariable logistic regression analysis to adjust the five covariates mentioned above. Spearman correlation was used to test association of the number of *APOBEC3A/B* allele (0,1,2) and continuous variables including genetic ancestry, COSMIC mutation signatures, and Ecotype signatures. For associations with FDR < 0.05, multivariable linear regression was used to further examine the association of a signature (continuous outcome variable) with *APOBEC3A/B* deletion (primary explanatory variable) adjusting for IA ancestry, age at diagnosis, tumor stage, PAM50 subtype, and TMB.

### Associations among the *APOBEC3A/B* deletion, TE expression, and CE10, and

We applied the REdiscoverTE pipeline ^40^ to RNA-seq FASTQ files from 720 Hispanic/Latina (H/L) tumor samples to quantify both gene and transposable element (TE) expression. This generated two expression matrices: gene-level counts for approximately 56,000 genes and intergenic TE-level counts for 1,052 TEs. Both matrices were normalized using the relative log expression (RLE) method implemented in the *calcNormFactors* function from the limma package ^69^. Normalization factors were computed based on total gene expression counts and applied to both gene and TE expression matrices. TE expression values were then transformed to log2 counts per million (log2CPM) with a prior count of 5.

To test our hypothesis that the *APOBEC3A/B* deletion may influence the CE10 immune ecotype through modulation of transposable element (TE) activity, we evaluated the association between the *APOBEC3A/B* deletion and TE expression by fitting multivariable linear regression models for each TE, with the *APOBEC3A/B* deletion coded as 0, 1, or 2 copies and adjusting for tumor purity, IA ancestry, TMB, and PAM50 subtype.. Secondly, we performed multivariable linear regression analyses to assess associations of CE10 and each TE individually, adjusting for the same set of covariates. Next, we applied LASSO regression to model CE10 as a function of all 1,052 TEs, adjusting for the same covariates, to identify a parsimonious set of TEs predictive of CE10. Model tuning was performed using 10-fold cross-validation, and the optimal penalty parameter was selected using the λ se criterion (the largest value of λ within one standard error of the minimum cross-validated error) to favor model sparsity and robustness.

## Data Availability

All data produced in the present study are available upon reasonable request to the authors

https://pubrepo.coh.org

## ACKNOWLEDGMENTS

This work was funded by the NCI (U54CA302452), NIMHD (R01MD021367) and the California Initiative to Advance Precision Medicine (OPR18111). Research reported in this publication included work performed in the City of Hope Integrative Genomics Core and Pathology Core supported by the NCI of the NIH under grant number P30CA033572. The content and views are solely the responsibility of the authors and should not be construed to represent the views of the NIH. The authors would like to acknowledge the work provided by the Leadership and Staff of the COH Center for Informatics, most notably Research Informatics, and the utilization of the POSEIDON data exploration, visualization, and analysis platform including the Honest Broker process. S.L. Neuhausen and some analyses were partially funded by the Morris and Horowitz Families Professorship.

